# A global assessment of dengue seasonality: Applying a novel, proportion-based method to case time series from 1990 to 2024

**DOI:** 10.64898/2026.07.13.26358002

**Authors:** Kishen Joshi, Katie M. Susong, Ahyoung Lim, Yang Liu, Oliver J. Brady

## Abstract

Dengue is a mosquito-borne, viral disease of increasing public health significance. Currently, most public health interventions target the vector, with efficacy dependent on timing within the season. Whilst seasonal profiles have been characterised in some endemic settings a global assessment is lacking. Here, we develop and apply a proportion-based measure of dengue seasonality to reported case time series from 1990 to 2024 across 106 countries and territories, the largest assessment of this phenomenon to date. We identify regional differences in seasonality such that every month of the year saw cases peak in at least one country or territory. Latitude was identified as influencing seasonality, with cases peaking between March and April in the southern hemisphere and July and October in the northern hemisphere. Equatorial locations displayed flat seasonality, and amplitude increased with distance from the equator. K-means clustering identified three seasonal profile types: two with pronounced seasonal outbreaks (with distinct peak timing and shape) and one with flatter, more endemic transmission. Peak month timing covaried among locations within the same seasonality cluster, with phase differences meaning that information on shifts in peak timing may be available several months in advance in some settings, of potential significance for prediction and intervention planning. Beyond aiding public health planning, identification of seasonal clusters suggests that information on dynamics in one location could be leveraged to improve forecasting power in others with similar seasonal dynamics.

**Author summary:** Understanding dengue seasonality is crucial for effective public health response. Most current interventions target the vector, with efficacy dependent on timing within the season. Previous work has shown that early implementation of interventions, targeting the vector when cases are increasing most rapidly rather than at their peak, leads to greater reduction of future incidence. Timing interventions in this way requires an understanding of the seasonal distribution of cases, and how consistent these patterns are between years. Despite this importance, seasonal profiles have only been characterised in a few endemic settings, leaving patterns in many at-risk locations poorly understood.

Here we address this gap by developing and applying a new, proportion-based measure of dengue seasonality to case time series from 1990 to 2024 across 106 countries and territories. Measurements of peak timing and its variability bear immediate utility for decision making regarding intervention timing by public health agencies. We also define a new temporal window for considering dynamics, the “dengue season”, which adjusts for phase differences in annual time series. Clustering of seasons identifies three seasonal patterns. Observations of concordant peak shifts amongst locations within the same cluster identifies relationships with potential for outbreak early warning and improving forecasting power.

## Introduction

Dengue is a viral disease transmitted primarily by *Aedes aegypti* and *Aedes albopictus* mosquitoes, of increasing public health significance (1). An estimated 390 million infections occur annually, with 96 million presenting symptomatically (2). Incidence has increased in recent times, with the World Health Organisation (WHO) estimating a 30-fold increase over the past 50 years (3). These increases are in part attributable to increased habitat permittivity for the vector, brought about by urbanization, globalization and climate change (4). This increased range increases the population at risk of infection (5). In 1990, an estimated 1.5 billion people lived in locations at risk of dengue transmission, approximately 30% of the global population (5). This has increased to an estimated 5.7 billion people, nearly 70% of the global population (6,7).

Dengue transmission intensity is dependent on vectorial capacity, which is influenced by a range of climatic factors. Temperature influences environmental suitability, vector lifecycle progression rate, population dynamics and biting rate, and extrinsic incubation period (8–18). Rainfall influences vector breeding site availability, via accumulating in containers which provides a habitat for oviposition, and subsequent larval and pupal stages (9,19). Changes in vector populations are observed throughout the year, in line with changes in climatic conditions. Changes in dengue transmission follow, with a lag accounting for time required for vector development and infectivity (8,20,21).

Transmission is also influenced by non-climatic drivers. In particular, human movement can influence timing of seasonal outbreaks by introducing dengue to locations without circulating virus (22–25). This bears significance in locations with climatic extremes unsuitable for year-round vector maintenance. Here, vector populations shrink when conditions are unsuitable and resurge when they are next suitable. In the absence of efficient vertical transmission the resurgent population contains little to no circulating virus (26). As such, seasonal outbreaks require re-introduction of the virus, either through transport of infected mosquitoes or movement of infected humans. Crucially, outbreak timing here depends on both the onset of the *Aedes*-suitable window and the timing of virus introduction, which can be after the start of the permissive window. Seasonal outbreaks with such characteristics are observed in emerging settings, including mainland China and Pakistan (23,25).

Population immunity has also been shown to modulate the influence of changes in vector capacity on transmission (27). Where there is a large pool of susceptible individuals, the effect of climate on transmission is more pronounced. Similarly, where there are few susceptible individuals, reduced climate forcing of transmission is observed. Susceptible pool size and serology are difficult to fully capture due to complexity of dengue immune responses. Dengue virus exists as four serotypes, with infection by a given serotype providing long-term immunity to that serotype and a short-term period of cross-protection against other serotypes (28). This mechanism generates a complex immune landscape, exacerbated by simultaneous circulation of multiple serotypes. Some studies have used serotype data to examine the effect of population immunity on dengue transmission, however the cost of serological data collection has prevented assessment on a global scale (29,30). As such, unobserved population immunity should be considered when developing methods to assess dengue transmission patterns.

The individual and combined effects of climatic and non-climatic drivers on transmission introduce an important feature of dengue incidence patterns, namely intra-annual variability whereby some-times of year experience higher incidence than others ^31^. Consistency in these patterns between years introduces the concept of dengue seasonality, where intra-annual differences in cases follow a regular pattern. Understanding this seasonality bears significance for planning public health interventions.

With an absence of antivirals, and the WHO only recommending vaccine use in high-transmission settings, most interventions target the vector (32–35). Many of these interventions involve reducing the density of containers representing potential oviposition sites (34). Crucially, efficacy of these interventions depends heavily on timing within the season. The WHO recommends earlier timing of interventions within the season, which has been shown to lead to greater reduction of later incidence (36–39). Timing interventions in this way requires understanding of the underlying dengue seasonality, as well as timing of the peak transmission month, and how variable it is between years.

Existing work considering temporal dynamics of dengue transmission confirms existence of seasonality, however more focus is placed on identifying relationships between transmission and climatic variables (40–53). Whilst understanding these relationships is important for forecasting system development, here it is at the detriment of improved understanding of seasonality (8,54). Considering this prior work, three key knowledge gaps remain. Firstly, the focus on variables with predictive power has led to reduced holistic understanding of dengue seasonality. Secondly, these studies focus on only a few at-risk locations, with patterns in many others poorly understood, leading to incomplete geographic understanding of seasonality. Thirdly, this focus on specific settings has reduced our ability to compare seasonal patterns between locations and extrapolate wider scale patterns. These latter two gaps are in part attributable to a lack of globally comprehensive data.

This study addresses these gaps by using a new, proportion-based metric to measure dengue seasonality in a scale-independent manner, facilitating comparison of patterns between years and locations independently of differences in total cases. We apply this method to dengue counts data drawn from OpenDengue (55), a recently released global dengue counts database, to provide a geographically-extensive assessment of dengue seasonality. Specifically, we measure facets of dengue seasonality with significance for intervention planning, including peak month timing; as well as identifying locations with similar seasonality across geographic space, bearing significance for prediction.

## Results

### Global seasonality of dengue

Global dengue seasonality was primarily shaped by case dynamics in South America and South-East Asia, which accounted for 49.6% (95% CI 45.1 – 53.8%) and 31.3% (95% CI 27.3 – 35.2%) of annual global cases respectively. Central America and South Asia follow, contributing to 10.3% (95% CI 8.9 – 11.7%) and 5.2% (95% CI 3.6 – 7.2%) of cases (Table 1). Global case numbers are highest in March and April, with 21.8% (95% CI 19.6 – 24.1%) of annual cases occurring during these months (Supplementary Table 1). This is primarily driven by South America, which contributes 74.4% (95% CI 69.5 – 78.9%) of the cases observed during the global peak (Fig. 1A, Table 2).

**Fig 1.**
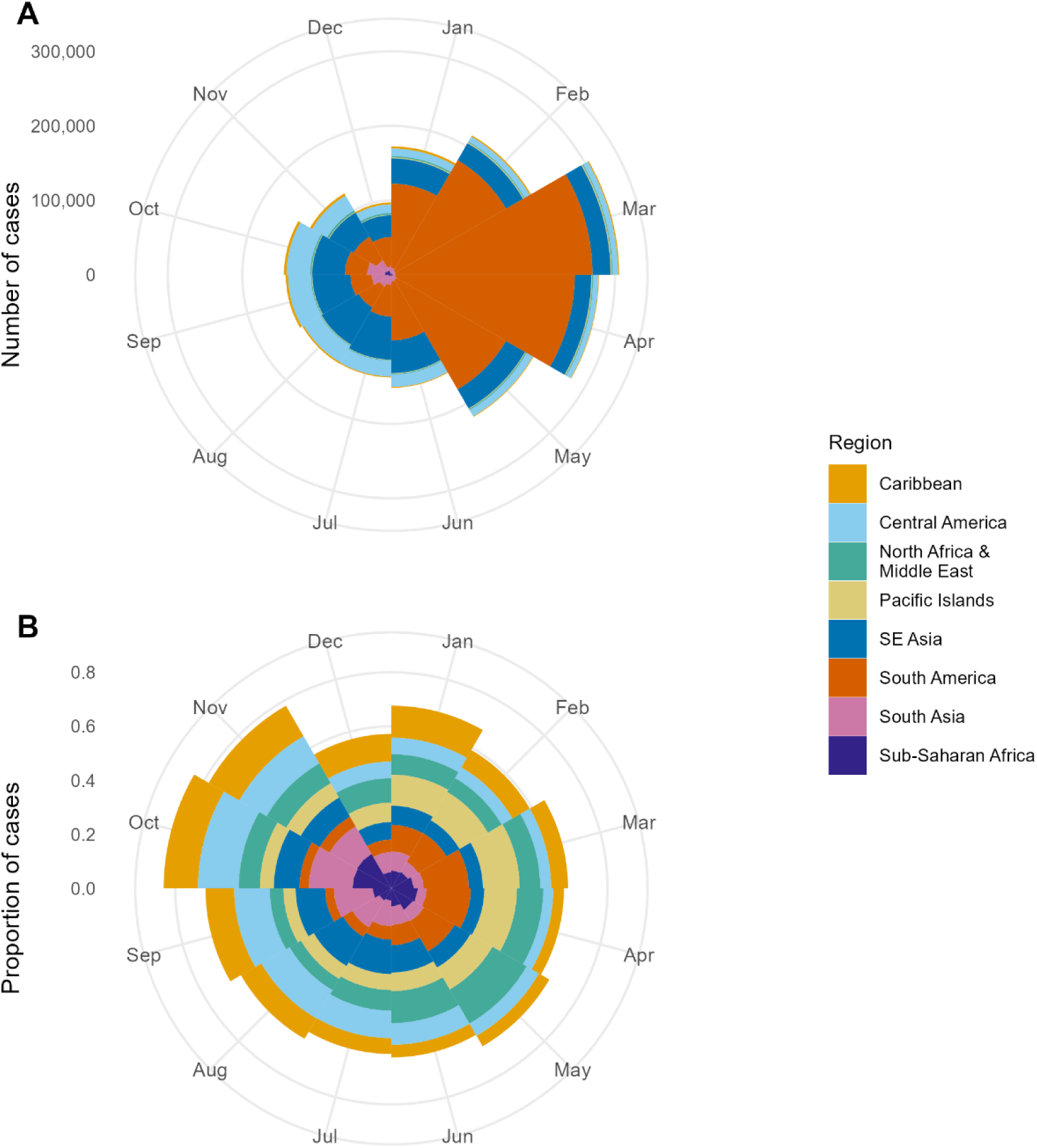
Radial bar plots of distribution of dengue case load throughout the year by region. Panel A shows average monthly raw case counts. Panel B shows average monthly proportion of annual case

**Table 1.** Percentage of annual global cases by region.

| **Region** | **Lower 95% CI** | **Average percentage of annual global cases** | **Upper 95% CI** |
| --- | --- | --- | --- |
| Caribbean | 1.6 | 2.2 | 2.9 |
| Central America | 8.9 | 10.3 | 11.7 |
| North Africa & Middle East | 0.3 | 0.5 | 0.8 |
| Pacific Islands | 0.5 | 0.7 | 1 |
| SE Asia | 27.3 | 31.3 | 35.2 |
| South America | 45.1 | 49.6 | 53.8 |
| South Asia | 3.6 | 5.2 | 7.2 |
| Sub-Saharan Africa | 0.5 | 0.8 | 1.3 |

**Table 2.** Percentage of cases at global peak contributed by each region.

| **Region** | **Lower 95% CI** | **Percentage of annual peak cases** | **Upper 95% CI** |
| --- | --- | --- | --- |
| Caribbean | 0.8 | 1.5 | 2.6 |
| Central America | 3.3 | 4.2 | 5.1 |
| North Africa & Middle East | 0.2 | 0.4 | 0.6 |
| Pacific Islands | 0.7 | 1 | 1.3 |
| SE Asia | 13.5 | 16.6 | 20 |
| South America | 69.5 | 74.4 | 78.9 |
| South Asia | 1.1 | 1.7 | 2.5 |
| Sub-Saharan Africa | 0.3 | 0.5 | 0.9 |

Large disparities exist in the number of dengue cases observed within each global region, making it difficult to compare seasonality between them (Tables 1 & 2, Fig. 1A, Supplementary Tables 2 & 3). The proportion-based metric used here increased our ability to make comparisons between different global regions with different population size and burden. This method shows that while global cases are highest between March and April, regional timing differs (Fig 1B, Table 3). Only 14% of countries and territories experienced peak cases during March and April. Instead, the largest proportion (18%) experienced peak cases during October, though every month of the year saw cases peak in at least one country or territory (Supplementary Table 4).

**Table 3.** Average dengue season peak by region.

| **Region** | **Lower 95% UI** | **Average peak** | **Upper 95% UI** |
| --- | --- | --- | --- |
| Caribbean | October | November | February |
| Central<br>America | July | September | November |
| North Africa &<br>Middle East | September | November | May |
| Pacific Islands | December | February | March |
| SE Asia | July | September | December |
| South America | February | April | July |
| South Asia | September | October | October |
| Sub-Saharan<br>Africa | October | October | November |

### Association between seasonality and latitude

Dengue seasonality varied with latitude. Cases peaked later in the year in the Northern hemisphere and earlier in the Southern hemisphere, with peak-to-trough distance increasing with distance from the equator (2.3% per degree, p < 0.001, Supplementary Fig. 1). Equatorial locations displayed minimal seasonality, with only small peak-to-trough distance observed (Fig. 2, Supplementary Table. 5). Statistically significant differences were observed between seasonality across all latitude bands, with effect size increasing when compared with more distal latitude bands (p < 0.001, Dirichlet regression likelihood ratio test, Supplementary Table 6). Across all latitude bands, the proportion of cases observed by month varied between years. Overall, months with greater average proportion of annual cases displayed greater between-year variability in proportion borne. This result indicates that while timing of high-risk months is consistent between years, the proportion of annual cases observed within these months is variable (Pearson correlation 0.71, Fig. 2, Supplementary Fig. 2).

**Fig 2.**
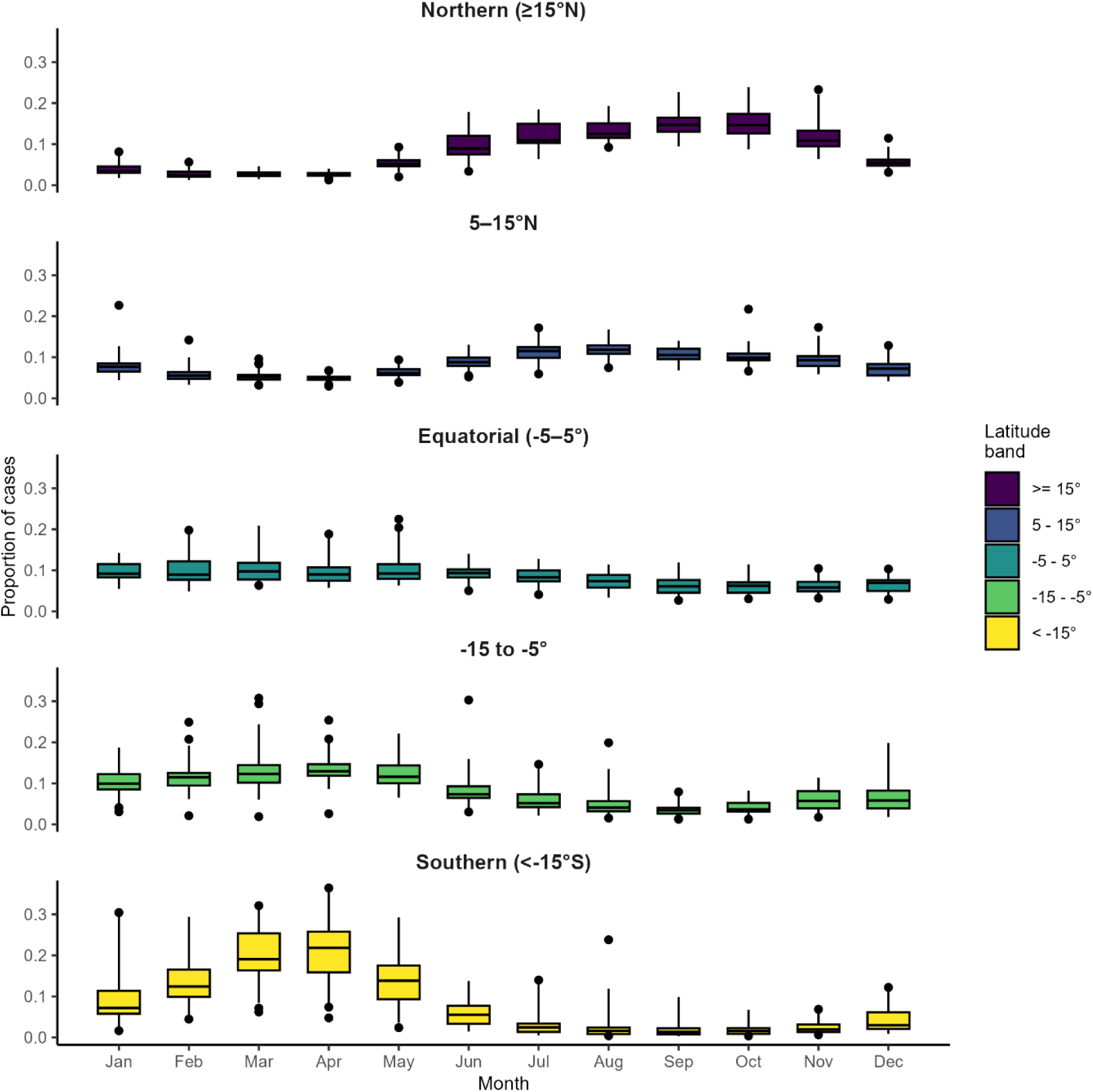
Boxplot showing distribution of monthly proportion of annual cases by latitude band.

### Peak month timing and variability

Peak month varies with latitude, becoming earlier with movement from North to South. Differences in timing are observed between locations at similar latitudes, notably between South Asia, where cases peak between September and October (Pakistan and India), and South-East Asia where cases peak between July and August (Myanmar and Thailand). Similar differences between locations at the same latitude were observed in South America. Inland locations overlapping with the Amazon rainforest had an earlier peak month timing, between January and February, compared to those closer to the coast where cases peaked between March and April (Fig. 3A).

**Fig 3.**
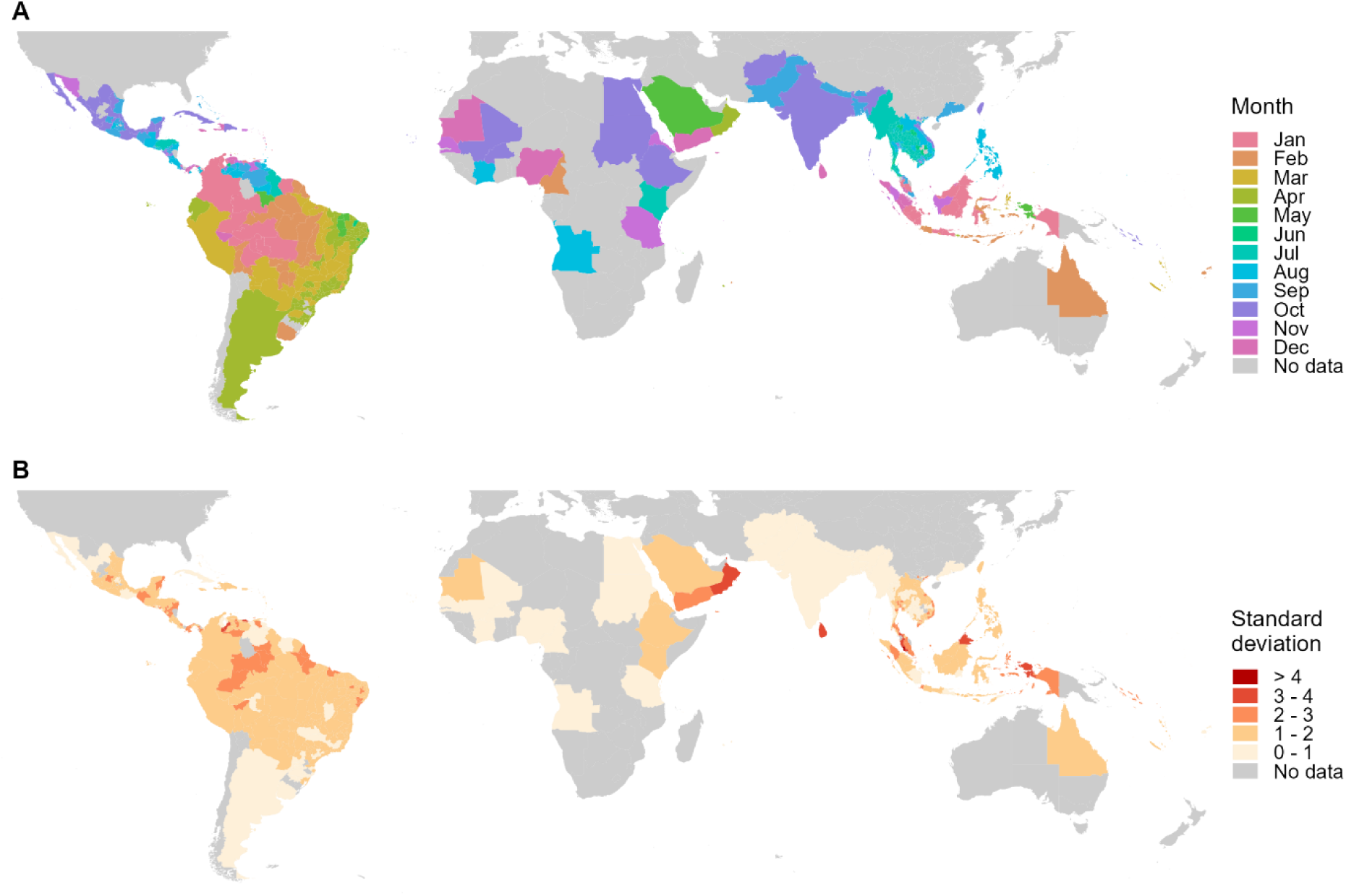
Dengue peak timing and between-year variability. Panel A shows peak month. Panel B shows variability of peak timing, measured using standard deviation of peak month number.

Peak month timing also varied between years. Timing was consistent in South Asia and Africa, with standard deviation of peak month timing below one month in most countries and territories. This contrasts with Central America, South America and South-East Asia where standard deviation of peak month timing was between one and two months for most locations. Parts of Venezuela, all the Middle East, Indonesia and Sri Lanka exceed this with standard deviation of peak month upwards of two months, reaching nearly four months in places (Fig. 3B, Supplementary Tables 7 & 8).

### Identifying clusters of seasonality

Dengue time series were re-aligned from calendar year to dengue season to identify locations with similar seasonal dynamics. The month with lowest average incidence was set as first (lowest) month of the dengue season. All clustering validation metrics used identified three clusters as optimal. (Table 4, Supplementary Fig. 3).

**Table 4.** Optimal number of clusters identified by each validation metric.

|  |  | Validation metric value |  |  |  |  |
| --- | --- | --- | --- | --- | --- | --- |
| Clustering validation metric | Optimal number of clusters | 2 | 3 | 4 | 5 | 6 |
| Silhouette score | Two or three | 0.23 | 0.19 | 0.16 | 0.16 | 0.15 |
| Elbow plot | Inconclusive | 65026.92 | 55057.09 | 49950.48 | 46428.43 | 43576.1 |
| Dunn score | Two, three or four | 0.02 | 0.02 | 0.01 | 0.01 | 0.01 |
| Calinski-Harabasz score | Two or three | 2632.22 | 2368.77 | 2046.96 | 1822.07 | 1670.62 |
| Davies Bouldin score | Three | 1.66 | 1.52 | 1.6 | 1.62 | 1.57 |
| Gap statistic | Three | 1.22 | 1.23 | 1.21 | 1.22 | 1.22 |

Three distinct types of dengue seasonality were identified. Cluster one and three had distinct high-transmission periods, with cases peaking in month eight and six of the season respectively. Cluster two also had a defined period of higher transmission however it was of lower amplitude, approximately 15.4% of the total seasonal cases compared to 25.4% for cluster one and 21.5% for cluster three (Fig. 4A, Supplementary Table 9, Supplementary Fig. 4).

**Fig 4.**
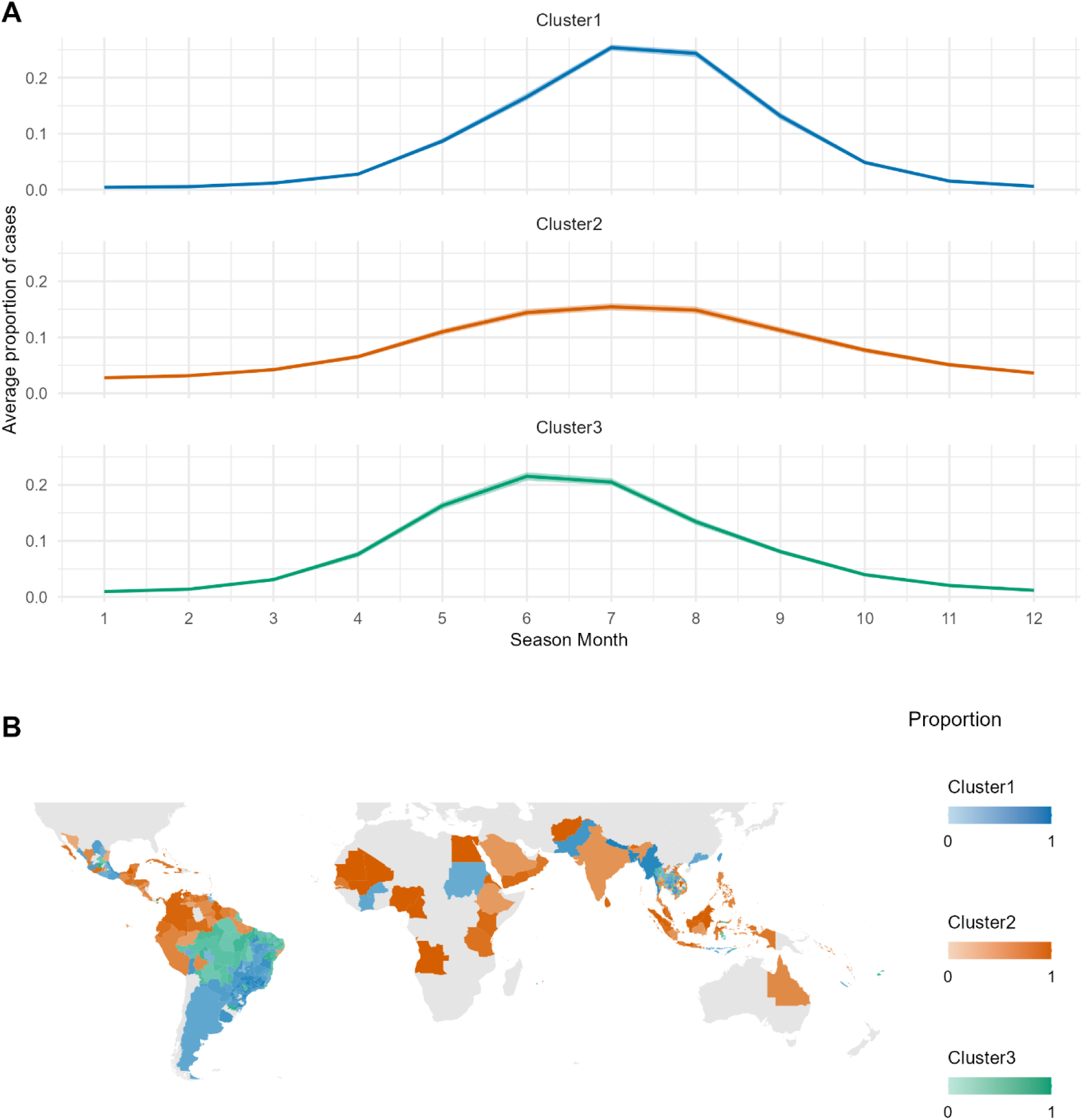
Seasonal profile clustering results.. Panel A shows average seasonal profiles by cluster. Panel B shows geographic distribution of clusters. Locations are coloured by cluster, with shade representing the proportion of times the location was allocated to the highest proportion cluster.

Central America, northern parts of South America, Africa and South Asia typically experienced cluster two seasonality. South-East Brazil, Argentina and South-East Asia typically experienced cluster one seasonality, while Northern Brazil typically experienced cluster three seasonality (Fig. 4B). Consistency of clustering was also of note. Locations following a cluster one pattern displayed no difference in proclivity for either cluster two or three when deviating (p > 0.05, Chi-squared). Cluster two locations displayed a greater proclivity to display a cluster three pattern when deviating (p < 0. 0.001, Chi-squared). Cluster three locations displayed a greater proclivity to display a cluster one pattern when deviating (p < 0.001, Chi-squared) (Fig. 4B, Supplementary Figs. 5 and 6).

### Implications of cluster allocation for prediction

To assess whether cluster allocation could support outbreak early warning, we examined whether season peak timing shifted consistently across locations within each cluster. For each location-season we calculated the shift in timing between the observed location peak and the cluster average. A shift of zero indicated complete agreement with the cluster average peak, whereas positive and negative shifts indicated later and earlier peaks, respectively. To assess whether timing shifts were consistent across locations we tested whether the median of the shift distribution differed significantly from zero. A median of zero would indicate no shift from the cluster average. In most seasons the peak month matched the cluster average (Supplementary Fig. 7). In some seasons however, consistent shifts were observed. Nine seasons in cluster one (25.7%), six in cluster two (17.1%), and eleven in cluster three (29.4%) displayed significant, cluster-wide shifts (Wilcox test, p < 0.05). Where shifts were observed, the peak most often occurred either one month earlier or later. For example, in cluster two the median peak shift in the 1999-2000 season was one month earlier, with 75 locations (65.8%) experiencing an earlier peak than expected (Supplementary Table 10).

These consistent shifts were identified in season-aligned data, which adjusted for phase differences in calendar-year-aligned time-series. As locations within the same cluster peak at different points in the calendar year, shifts observed in countries and territories with earlier peaks may support outbreak early warning for those with later peaks. Dengue seasons typically cross the calendar year, with locations with an earlier seasonal cycle peaking in the latter half of the calendar year, and those with a later seasonal cycle peaking in the first half of the next calendar year. In cluster one, peak shifts in early-season locations in South and SE Asia may anticipate shifts in South America (Fig. 5A, 5D). In cluster two, peak shifts in locations in South and SE Asia, Sub-Saharan Africa, the Caribbean and Central America may anticipate shifts in South America and the Pacific Islands (Fig. 5B, 5D). In cluster three, peak shifts in locations in SE Asia may anticipate peak shifts in South America (Fig. 5C, 5D).

**Figure 5.**
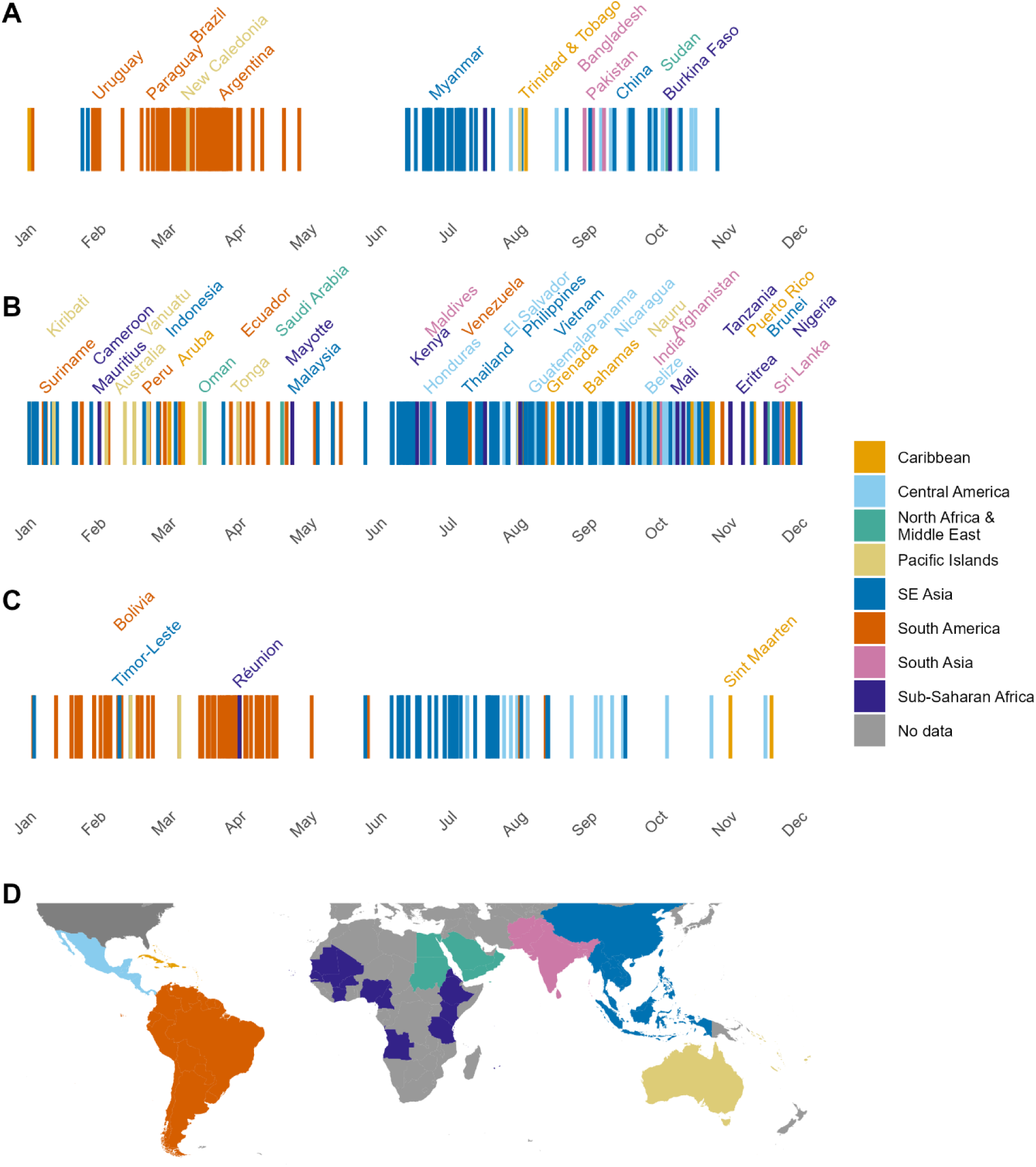
Geographic gradient in peak timing by cluster. Panels A, B and C show barcode plots of peak timing by location in cluster one, two and three respectively. Countries and territories are coloured by region. Panel D shows a map of regional grouping.

## Discussion

This study used recently released global dengue case count databases to provide the first global assessment of dengue seasonality, incorporating a dataset of over 100 million cases from 106 countries and territories. We quantified average seasonality and between-year consistency, revealing seasonal variability along a latitudinal gradient. Seasonality became more pronounced with movement from the equator, with Northern and Southern hemispheres displaying out-of-phase seasonality; as expected considering austral and boreal season timing (56,57). After adjusting for these phase differences, we identified three main seasonality patterns, with countries and territories in different parts of the world progressing through similar seasonal dynamics at different times of year. In seasons when the observed peak shifted from the cluster average, the direction of shift was concordant across locations within the same cluster.

These results have direct implications for intervention planning, outbreak early warning and short to medium term forecasting. Most current interventions target the vector, with efficacy dependent on timing within the season. Earlier implementation is associated with greater reduction of future incidence (36–38). Our quantification of peak timing and its between-year variability provides public health agencies with empirical guidance for identifying optimal timing for implementing vector control measures (Fig. 3, Supplementary Table 7). Identification of shared shifts in peak timing across clusters aids this planning. Concordant peak shifts among locations with the same seasonality pattern allow use of seasonal information from one location for inference in another. Moreover, grouping of early-and late-season locations identifies geographically distal regions with lagged seasonal dynamics, providing a more holistic perspective on likely shifts in peak timing. The geographic relationships identified (Fig. 5) can be leveraged to inform intervention planning and early warning.

This cluster-wise synchrony extends beyond intervention planning to forecasting system development. Dynamics in one location could improve forecasting power in other locations displaying the same seasonality pattern. Synchrony in dengue dynamics has already been observed across South America (58) and South-East Asia (59), identifying relationships of potential utility for prediction. Consistently, dengue forecasting systems in mainland China incorporate information on transmission from other South-East Asian countries (60). Our clustering approach identifies groups of geographically distant countries and territories with shared predictive power beyond these regional scales. Incorporating temporal dynamics of these locations into forecasting systems may improve prediction in the cluster as a whole. Quantification of seasonal variability complements this, providing information on ease of forecast-ability. Supplementary Table 8 provides a ranked list of countries and territories ordered by consistency of seasonality, identifying potential candidates for forecasting system development efforts.

Beyond operational applications, these observed patterns reveal fundamental features of global dengue epidemiology. Identification of a latitudinal gradient in peak season timing extends prior observations of regional synchrony to a global scale (58,59). Peak transmission months are observed earlier in the year in the Southern hemisphere, becoming later with movement northwards. Adjusting for these phase differences facilitated combination of seasonal data across all locations. Subsequent clustering analysis identified three common types of seasonality. Two seasonal types display defined seasonal outbreaks with little transmission outside this window. The third type exhibits cases year-round with reduced within-year variability. While further work is required to identify drivers of these patterns, we posit that seasonal changes in climate are of importance, via their influence on *Aedes* vectorial capacity (9–18,56,61). Considering their geographic position, locations with defined outbreaks likely experience distinct climatic seasons with defined periods of *Aedes* climatic suitability. Similarly, locations with year-round cases likely experience less well-defined climatic seasons and reduced variability in *Aedes* climatic suitability. Observations of concordant shifts in seasonal timing are suggestive of global climatic systems, for example the El Niño–Southern Oscillation, influencing the behaviour of local climatic drivers (62–66). Influence of these far-reaching climate systems could provide insights into a unified mechanism controlling cluster-wise synchrony in local dengue transmission dynamics.

While climatic suitability shapes baseline seasonal patterns, between-year variability in these patterns likely reflects influence of non-climatic drivers, particularly population immunity and spatial connectedness. Both can affect seasonality via their influence on the size and composition of the susceptible pool. Seasonal epidemic magnitude is constrained by the availability of susceptible individuals (27). Spatial connectedness may shape serotype and strain-level susceptibility by facilitating introduction of novel lineages, contributing to viral mixing and population turnover to potentially prevent build-up of immunity. As such, between-year changes in susceptible pool size and serological composition may help explain variability in peak amplitude and seasonal pattern.

Several data limitations warrant consideration when interpreting these results. Firstly, variable spatial resolution of reporting reduces our ability to assess heterogeneities in seasonality in countries and territories with large geographic areas spanning climactic and ecological zones, most notably in India. The assessment of seasonal variation as a function of latitude is potentially affected by this spatial aggregation of counts data across large areas. Taking the at-risk population weighted rather than geometric centroid helped assign locations to the most suitable latitude band; however, highly populous settings may still have disproportionately influenced the results.

Potential within-season differences in diagnoses represents another limitation. At the beginning of the dengue outbreak, dengue cases may be more likely mis-diagnosed due to lack of established awareness amongst clinicians that the dengue season is starting. Similarly, as the outbreak progresses other febrile illnesses may be wrongly diagnosed as dengue, as the clinical staff will have likely encountered more dengue cases in the recent past. This reliance on clinical diagnosis introduces potential risk of systematic bias at the data generation process level, potentially affecting this work.

Despite these limitations, these results highlight several exciting opportunities for future research. This work provides the largest-scale assessment of dengue seasonality to date, introducing a novel, proportion-based approach to measuring seasonality, of potential utility for other seasonal diseases. Identification of three underlying patterns of dengue, and correlated peak shifts within these clusters, bears significance for improving understanding of dengue epidemiology and potential to improve predictive power of existing systems. Future work to contextualise these cluster-wise relationships between climate and dengue transmission is critical for anticipating future changes in seasonality and mounting effective public health interventions.

## Materials and Methods

### Defining region

Regions were defined by adapting the World Bank definitions. Regional definitions were maintained for Sub-Saharan Africa, Middle East and North Africa, and South Asia. The Latin America and Caribbean region was split into Caribbean, Central America and South America. The East Asia and Pacific region was split into South-East Asia and Pacific Islands (69) (Appendix 3).

### Dengue case counts data sources

All case count data were obtained from the OpenDengue project. This project compiles historical dengue case data reported by Ministries of Health, WHO, and other dengue surveillance data platforms such as Project Tycho (55,67). The raw OpenDengue data ranges between 1990 and 2025, however is geographically and temporally incomplete within this period. As this analysis requires a complete dataset, a gap-filled version developed in a parallel analysis was used.

This previous analysis:

1. Compiled data for 143 countries and territories in total, including Africa and the Middle East for which data are under-represented.
2. Imputed values for missing months.
3. Disaggregated years with only annual totals into monthly counts.
4. Assigned zeros to years with no dengue reports in predefined emerging epidemic settings, where sustained endemic circulation was not expected and the absence of reports was more likely to reflect true absence of local cases than missing reporting.

Most of the resultant dataset was comprised of observed and zero-assigned data; most gaps in the original data arose from years reported only as annual totals. Monthly values for these annually aggregated years were obtained from temporal downscaling using a scaled-INLA model with a hierarchical seasonal structure. This formulation allowed places with sparse data to draw strength from patterns both globally and within the same latitude band. Full methodological details are available in the publication (68).

For finer resolution analyses, we used the OpenDengue highest spatial resolution dataset directly. Case counts data for 25 countries and territories at the first administrative level, and 10 countries and territories at the second administrative level was obtained here. Data at the second administrative level was aggregated to the first administrative level. Data at the second administrative level in Brazil was aggregated by mesoregion (Appendix 1).

After processing, exclusion criteria were applied (Appendices 2 & 3). Any years with fewer than five cases per month on average were removed, as their sporadic transmission pattern prevents assessment of underlying seasonality. Any locations with fewer than three years of data remaining were excluded for the same reasons. After processing and exclusion criteria application, data was available for 106 countries and territories, including 13 countries and territories with finer-resolution data.

### Defining latitude bands

Raster population data for 2025 was downloaded from Global Human Settlement Layers (GHSL), at 1×1 km resolution (70). An arbovirus risk map obtained from previous modelling work was overlaid on the population raster (6). This identified raster cells containing populations living in transmission-permissive locations. Polygons for each target spatial unit were overlaid on the at-risk population raster. The at-risk population weighted centre of the shape vector was identified. Locations were allocated into latitude bands using this weighted centre. A map of location allocation by latitude band is included in Appendix 4.

### Quantifying temporal distribution of cases

Two temporal reference periods were defined, calendar year and dengue season. Calendar year aligned data provided an absolute temporal window for comparison of dynamics. Dengue season aims to consider the shape of the underlying seasonality, adjusting for differences in phase. As such, seasons were defined as lasting 12 months, starting on the month with lowest average case counts (Appendix 5).

Taking proportion of cases occurring each month by calendar year or dengue season normalised differences in case counts between years and seasons. This rescaling facilitated comparison between time periods and locations with different case counts. Cm refers to the number of cases observed in month m. Pm refers to the proportion of annual or seasonal cases observed in month m.

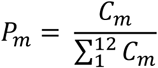

Taking the average of these monthly proportions returned an average profile for calendar year or season.

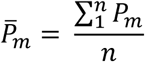

To measure consistency of the average seasonal profile, the Root Mean Squared Error (RMSE) of the proportions was calculated.

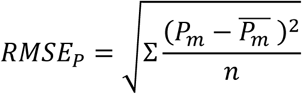

### Relationship between latitude and seasonality

#### Latitude and peak-to-trough distance

Relationship between latitude and peak-to-trough distance was assessed using a generalized linear model with Gamma-distributed errors and a log link. Let *Y_i_* denote the peak-to-trough distance for observation *i*, and *L_i_* = ∣ Latitude*_i_* ∣. We assumed:

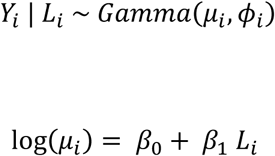

#### Differences in seasonality between latitude bands

Significance of differences in seasonality between latitude bands was assessed using Dirichlet regression. For each pair of latitude bands, we constructed annual seasonal compositions Y***_i_*** = (*Y_i_*_1_,…,*Y_i_*_12_), where Y*_im_* is the proportion of annual cases occurring in month m. We then fitted a Dirichlet regression model for each pair of latitude bands:

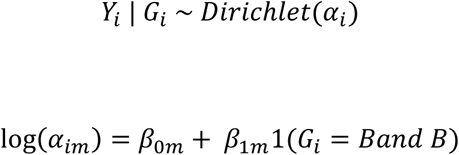

A null model assuming common seasonality across latitude bands was also fit:

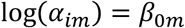

Models were compared using likelihood ratio tests, which follow a chi-square distribution, allowing calculation of p-values indicative of significant differences in seasonality between bands.

#### Magnitude of differences in seasonality between latitude bands

Magnitude of the difference in seasonality of latitude bands was assessed using Jensen-Shannon (JS) distance. P and Q are monthly proportion vectors, and Kullback-Leibler divergence (KL) is computed elementwise across all months.

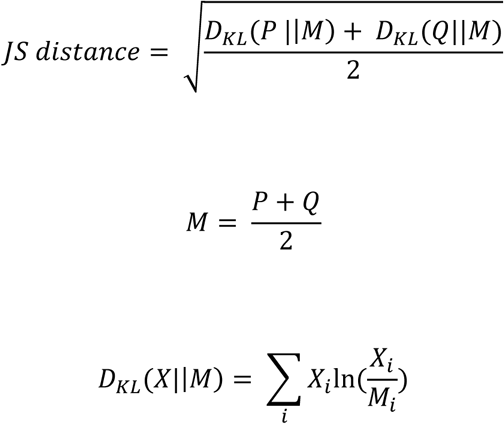

#### Identifying calendar year peak and between-year variability

Annual peak month was identified as the calendar month with most case counts. Where multiple months within the same year experienced equal cases, peak month was defined as circular mean of month number. Circularising peak month accounted for the underlying cyclical structure, whereby January (month one) is closer to December (month twelve) than to April (month four). Average peak month was then calculated across all years using a similar circular mean. Between-year variability in peak timing was calculated using circular standard deviation of peak month. Uncertainty intervals for regional peak month timing were also calculated in a manner consistent with the circular nature of the data. Here, 95% uncertainty intervals were identified as the edges of the arc within which 95% of peak month data points were observed. The circular R package (V0.5-1) was used for computation of circular statistics.

### Identifying clusters of seasonality

K-means clustering was applied to season aligned data to identify patterns of seasonality. Isometric log ratio (ILR) transformation was used to address incompatibility between the sum-to-one constraint of proportional data and Euclidean distance (Appendix 6). Following transformation, clustering was run 100 times, using a maximum of 100 iterations, for the k means parameter in the range one to ten. An elbow plot was generated to identify optimal number of clusters. Five other metrics were calculated for each number of clusters tested to aid decision making: Calinski-Harabasz score, Gap statistic, Silhouette score, Dunn score, and Davies Bouldin score. These metrics assess different aspects of clustering efficiency to indicate the optimal number of clusters (Appendix 7).

Overall, the elbow plot, Calinski-Harabasz score and Gap statistic assess within-cluster dispersion and the Davies-Bouldin index, and Silhouette and Dunn scores measure cluster separation. The number of clusters identified as consensus from these metrics was selected as optimal. Where no consensus was observed, candidates for optimal number of clusters were identified from the diagnostic plots. A visual assessment of results for each of these values was then performed to determine the optimal number of clusters to proceed with.

Once the desired number of clusters had been identified, kmeans time series clustering using 1000 runs with distinct starting positions was performed on all seasonal data. The proportion of times that each season was placed into each cluster was calculated. Seasons were placed into the cluster they were allocated the highest proportion of runs. Locations were placed into the cluster the highest proportion of their seasons were allocated to. When identifying the cluster average seasonal profile, calculations were performed in logit space to account for [0,1] bounds.

Cluster preference of seasons when deviating from the primary pattern was assessed using a chi-square test. Number of seasons falling into the non-primary cluster was counted for each secondary cluster. Expected number of counts was set to half the number of deviating seasons, indicative of no preference.

Likelihood of peak month to deviate from the average was also assessed. Difference between observed peak month and the cluster average was calculated for each location by season. The significance of the difference between the resulting distribution and the zero-reference distribution was assessed using a Wilcox test.

### Implications of cluster allocation for prediction

To assess utility of cluster allocation for peak shift prediction, we first identified the month with highest cases for each location and season, hereafter termed the “peak month”. Taking the average of these peak month values across locations and seasons within the same seasonality cluster provided a mean peak timing by cluster. We then calculated the difference in observed peak month for each location and season from the cluster peak, hereafter termed “peak shift”. A peak shift of zero indicates perfect agreement with the cluster-level expectation, while negative and positive values indicate earlier and later peaks, respectively.

We then assessed whether peak shifts were centred around zero, as would be expected if clustering captured peak timing perfectly. The distribution of peak shifts was evaluated for normality using the Shapiro–Wilk test. As the distributions departed from normality, we used a Wilcoxon signed-rank test to assess whether the median peak shift differed from zero. Seasons whose median peak shift departed from zero were considered to shift from the cluster peak.

## Data availability

High spatial resolution dengue counts data was obtained from OpenDengue (https://opendengue.org/data.html). Dengue counts data for nations and territories were obtained from a gap-filled version of the OpenDengue national dataset developed in a parallel analysis (68). Population raster data were obtained from the Global Human Settlement Layers database (https://human-settlement.emergency.copernicus.eu/download.php?ds=pop). Information on nested administrative levels in Brazil was obtained from publicly available resources provided by the Brazil Institute of Geography and Statistics (https://www.ibge.gov.br/en/geosciences/territorial-organization/regional-division/23708-brazilian-territorial-division.html?=&t=downloads). Dengue risk maps were obtained from the publicly available GitHub repository: https://github.com/ahyoung-lim/Arbo_riskmaps_public (6).

All data used in this study are publicly available from the sources listed above, and are available online at in the study Zenodo repository: 10.5281/zenodo.19033464.

## Code availability

All analyses were performed in the computing software R (version 4.4.1) (71) and RStudio (v2023.12.1)(72). Key R packages include: tidyverse (v2.0.0) (73), bios2mds (v1.2.4) (74), fpc (v2.2-13) (75), cluster (v2.1.6) (76), clusterCrit (v1.3.0) (77).

All code used to generate the results reported in this study is publicly available online in the study GitHub repository (https://github.com/KishenJoshi/global_dengue_seasonality).

## Appendices

### Appendix 1: High spatial resolution data processing

The OpenDengue highest spatial resolution dataset contains data at the first and second administrative level, with spatial resolution varying between countries and territories. Linear interpolation was applied to each spatial unit (Appendix 2). Dengue counts reported at the second administrative level were aggregated to the first administrative level to ease interpretability and reduce noise. Data was available for 2007 locations at the second administrative level, contained within 165 spatial units at the first administrative level. To ensure robust aggregation, any years for administrative level one units with incomplete data at the second administrative level were removed. After application of this exclusion criterion, enough second administrative level data remained to aggregate data for 40 units at the first administrative level. This exclusion criterion was not applied to locations with counts data available at the second administrative level at the edge of the dengue distribution, namely Argentina and Mexico. Missing data at the second administrative level in these countries is assumed to relate to a lack of cases rather than true missing data.

In Brazil, data at the second administrative level was aggregated by mesoregion. Mesoregions in Brazil represent an intermediate level of organization between the first and second administrative levels. Brazil has 27 states, the first administrative level, however their large size and range across multiple latitudes means they include many environmental and climatic contexts. Performing analysis by mesoregion instead of state captures intra-state variation while avoiding the potential noise.

OpenDengue draws Brazil counts data from the notifiable diseases reporting system, SINAN, which reports data at the second administrative level. Crucially, SINAN omits data for units with zero counts within a year. All available data at this resolution was therefore aggregated by mesoregion in Brazil, as data gaps are assumed to relate to lack of cases, rather than true missing data.

Following aggregation, countries and territories without at least 50% of higher resolution units remaining following processing were excluded from the high spatial resolution analysis. This 50% exclusion criterion was not applied to countries at edges of the permissive dengue range, namely Australia and mainland China. Instead, high spatial resolution data for these locations was retained, as lack of data likely related to lack of cases instead of gaps in reporting. Once this data was prepared any years with fewer than five cases per month on average were removed, as their sporadic transmission pattern prevents assessment of underlying seasonality. Finally, any locations with less than three years of data remaining were excluded.

### Appendix 2: Dengue counts data processing

The same data processing steps were applied to each location, country and territory. Linear interpolation was applied to gaps no longer than one month or four weeks, using data points on each side to fill the intervening period. Any years with larger gaps were excluded. Weekly OpenDengue data was aggregated by month, generating a dataset with consistent monthly reporting. For weeks spanning two months, the average number of daily cases within that week was calculated. Daily case counts were then allocated to their respective months.

### Appendix 3: Regional classification

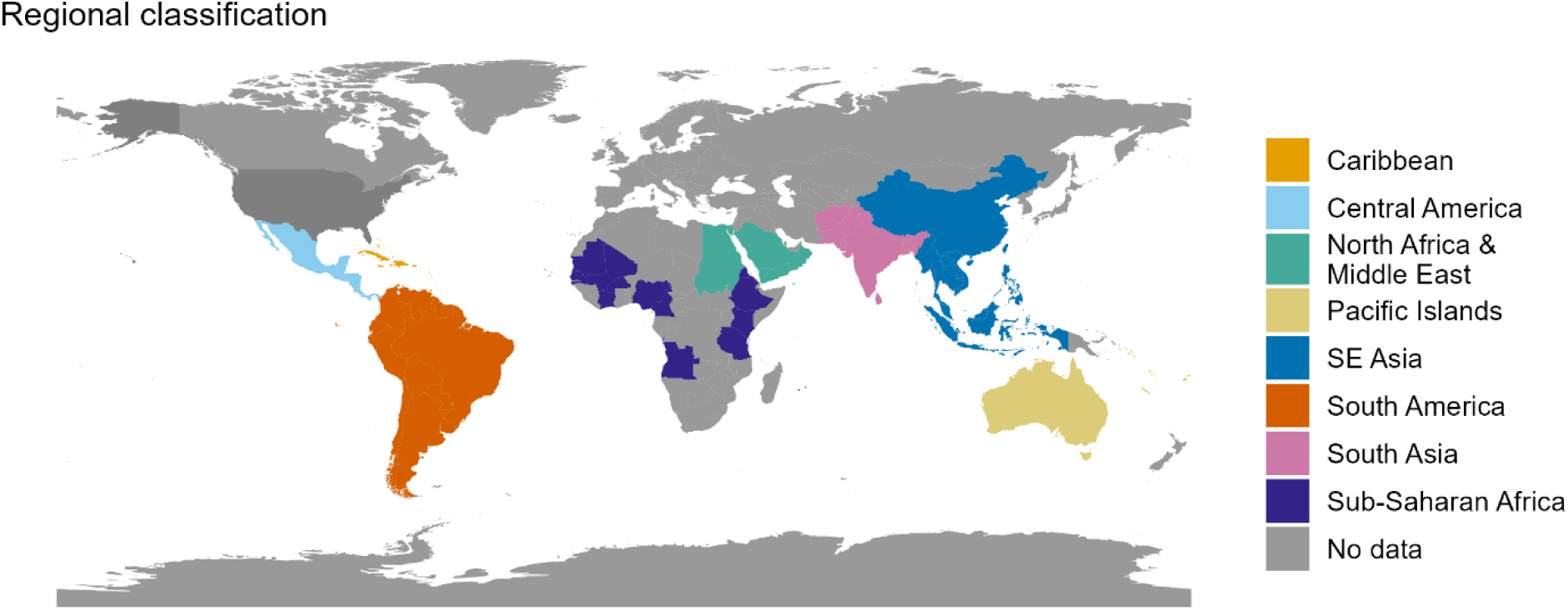

### Appendix 4: Latitude band allocation

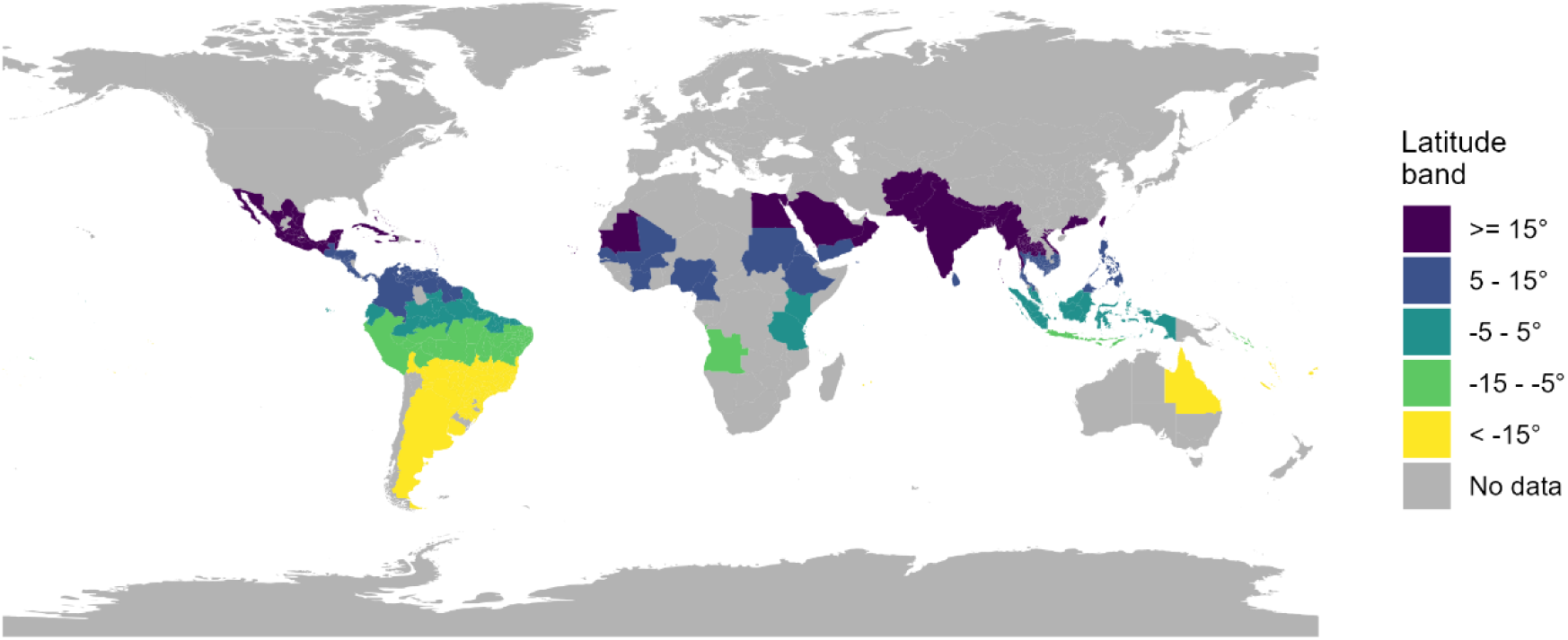

### Appendix 5: Seasonal alignment

Dengue season aims to consider the shape of the underlying seasonality, adjusting for differences in phase. Start of the season was defined as calendar month with lowest case count. Where counts were equal across multiple months within the same year, circular average of these months was taken. In the specific case where lowest counts for a year occurred in two months opposite each other on the calendar, e.g. June and December, a circular mean is incalculable. In these cases, the mean was taken. This yields two options, in this example March and September, months three and nine respectively. The option that maximised distance from the peak month for that year was selected. Average low month was then calculated across all years using a circular mean. This was set as month one of the season, and all months re-aligned accordingly.

Similar exclusion criteria were applied to dengue season as calendar year. Namely, any seasons with fewer than 5 cases per month on average, or 60 cases per season were excluded. Following application of this exclusion criterion, any locations with fewer than three seasons remaining were also excluded.

### Appendix 6: ILR transformation

K-means clustering uses Euclidean distance to measure separation of clusters and optimise accordingly. Euclidean distance however is not viable for assessment of proportional data due to the sum-to-one constraint. Isometric-log-ratio (ILR) transformation was used to convert proportional data into a form compatible with Euclidean distance.

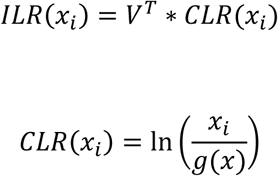

The centred log-ratio (CLR) transformation takes the logarithm of the ratio of each part to the geometric mean of all parts. ILR transformation multiples the CLR by a matrix representing the orthonormal basis of the CLR plane, returning uncorrelated orthogonal coordinates within a Euclidean space. This resultant linearly independent data is suitable for use within k-means clustering.

### Appendix 7: Clustering summary metrics

An elbow plot was generated showing the within-cluster sum-of-squares (WCSS) as a function of cluster number. WCSS decreases with increasing cluster number. Plotting WCSS against cluster number allows identification of the point where WCSS declines less steeply with increasing number of clusters, indicating diminishing returns. Number of clusters corresponding to this point, the elbow, is identified as optimal.

The Calinski-Harabasz score was calculated as the ratio of between-cluster sum-of-squares (BCSS) to WCSS multiplied by the ratio of the number of data points (N) minus number of clusters (k) to the number of clusters minus one. The score ranges from zero to infinity, with a higher value indicating better clustering.

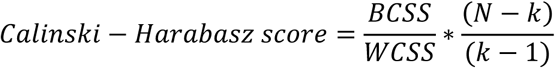

The Gap Statistic compares the within-cluster dispersion of the data to that of a null reference distribution. B reference datasets are generated from a random uniform distribution across the same range as the original data. Each of these is clustered for the same range of k values and WCSS calculated for each k. The gap statistic is calculated as the difference between the average of the logarithm of the WCSS of the reference datasets and the WCSS of the original data. The optimal number of clusters is selected as the smallest value of k that maximises the gap statistic, indicating the largest displacement from the uniform reference distribution. The gap statistic plot highlights the value of k whose gap statistic value is no more than one standard error from the gap statistic of the first local maximum, the default of the clusGap function from the cluster R package.

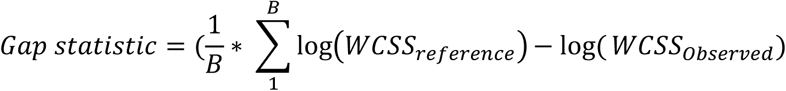

The Silhouette coefficient is calculated for each observation in the dataset by first identifying the average distance between that observation and other observations within the same cluster (ai) and the minimum distance between that observation and all other observations (bi). The difference between bi and ai is divided by the maximum of bi and ai. The Silhouette score averages all Silhouette coefficients across the entire dataset. The score ranges between-1 to +1, where +1 indicates strong similarity to observations within the same cluster and-1 indicates strong similarity to observations within another cluster.

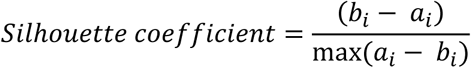

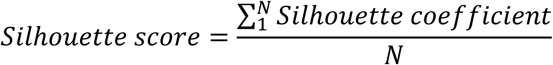

The Dunn score takes the ratio of the minimum separation between observations from different clusters to the maximum diameter across all clusters. Values range from 0 to infinity, with higher values associated with better clustering.

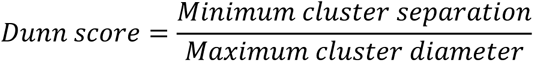

The Davies-Bouldin score takes the average similarity between each cluster and its most similar cluster, with similarity captured using Euclidean distance between cluster centres.

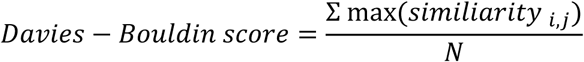

### Appendix 8: Packages and shapefiles

Analyses were performed using R version 4.4.1.

| Package | Version | Use |
| --- | --- | --- |
| bios2mds | V1.2.4 | Clustering |
| fpc | V2.2-13 | Calinski-Harabasz and Dunn scores |
| cluster | V2.1.6 | Silhouette score and Gap statistic |
| clusterCrit | V1.3.0 | Davies-Bouldin index |
| rnaturalearth | V1.0.1 | Shapefiles |
| geobr | V1.9.1 | Shapefiles for Brazil mesoregions |
| countrycode | V1.6.0 | ISO3 codes |

Shapefiles at the second administrative level were obtained from the Global Administrative Unit Layers (GAUL) dataset, implemented by the UN Food and Agriculture Organisation (FAO) within the CountrySTAT and Agricultural Market Information System (AMIS) projects.

