## Supplementary figures and images for "A global assessment of dengue seasonality: Applying a novel, proportion-based method to case time series from 1990 to 2024"

### Supplementary Figure 1

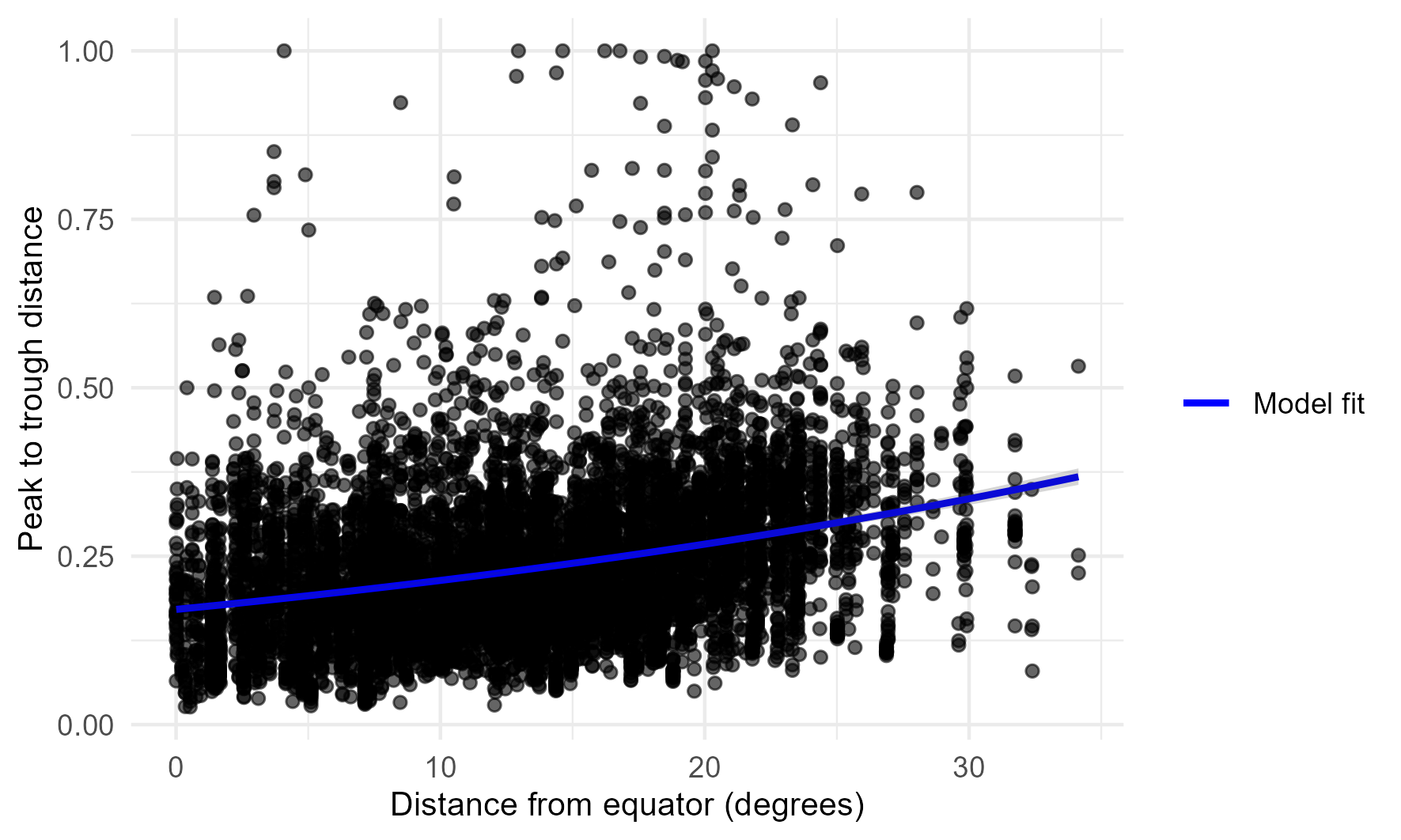

### Supplementary Figure 2

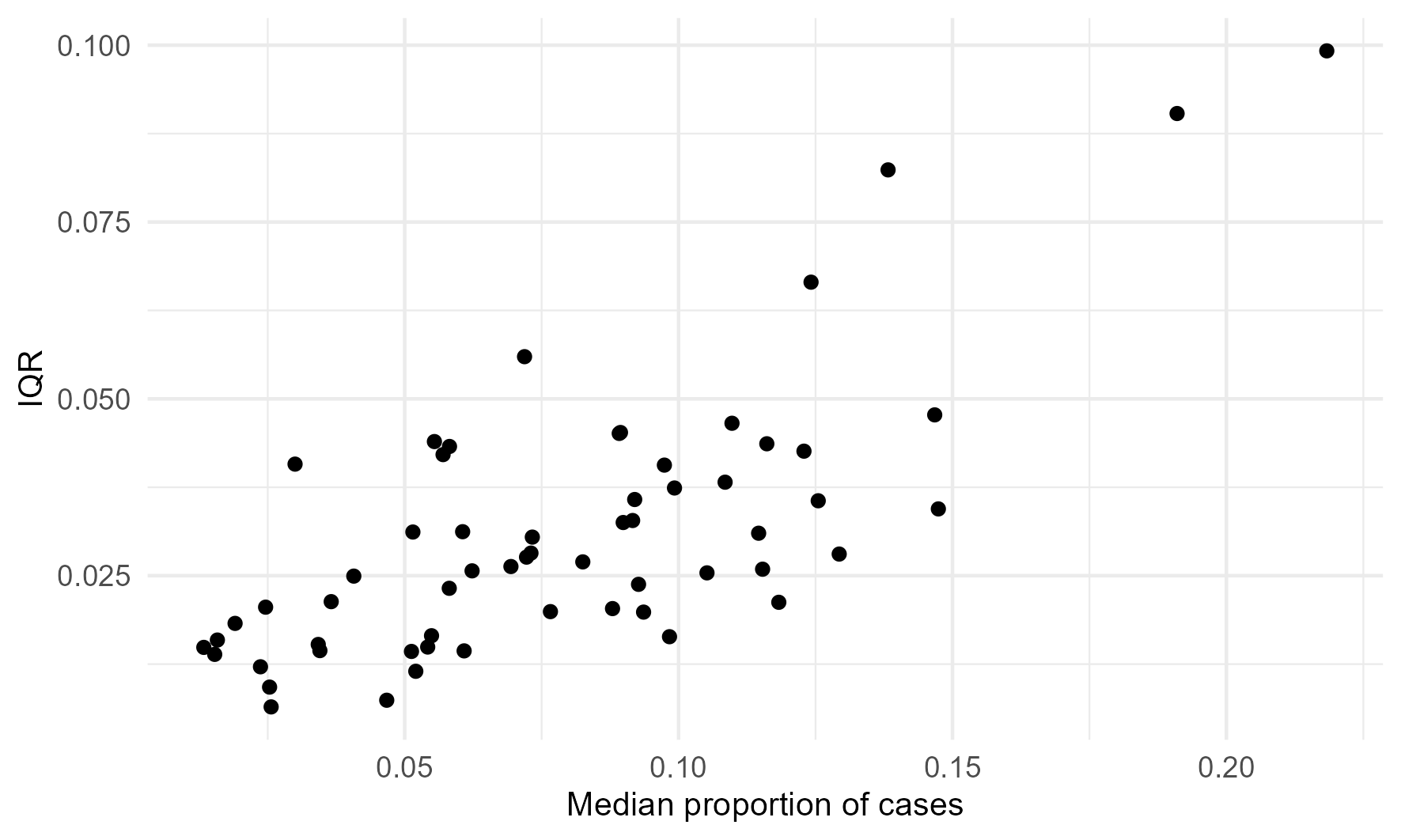

### Supplementary Figure 3

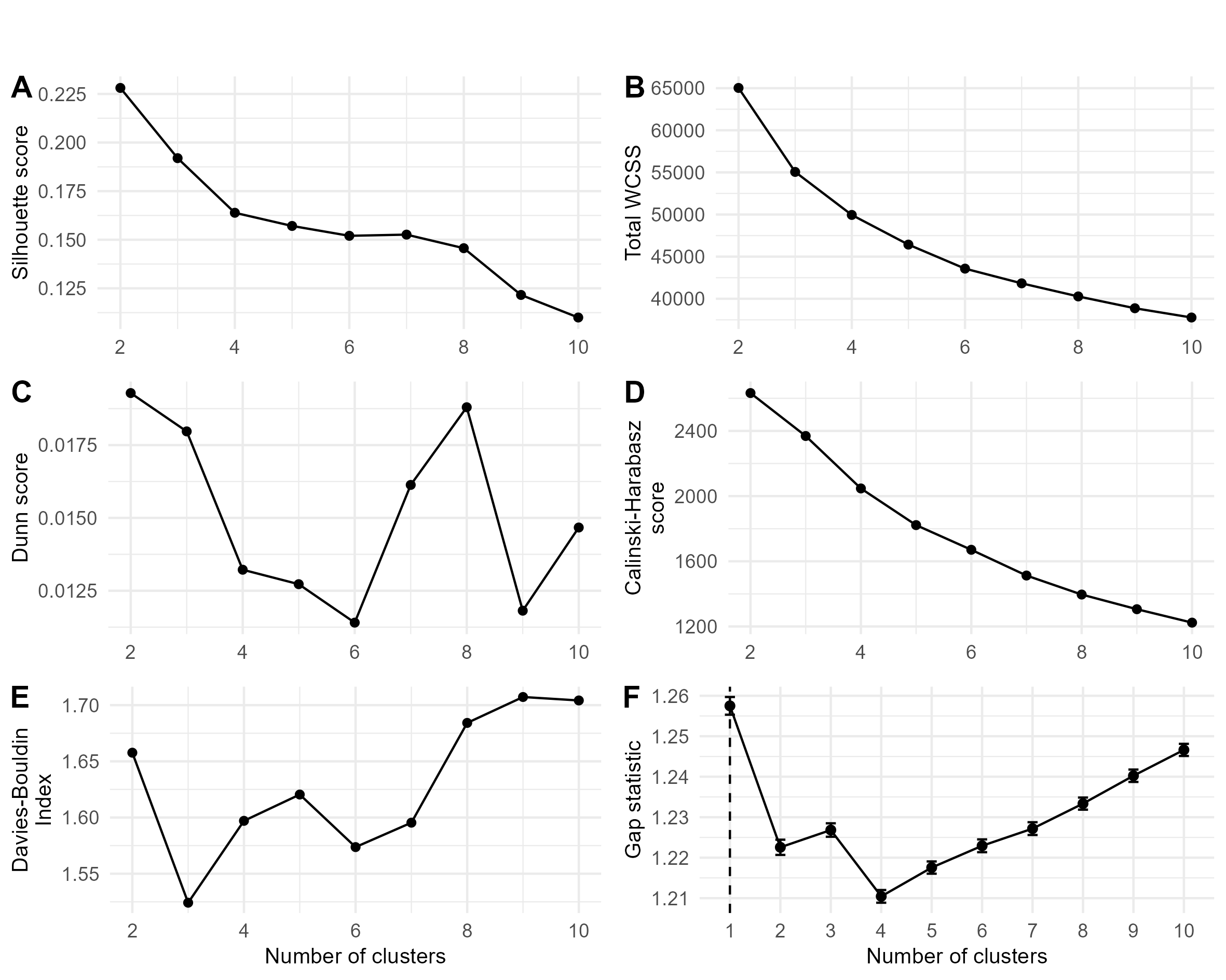

### Supplementary Figure 4

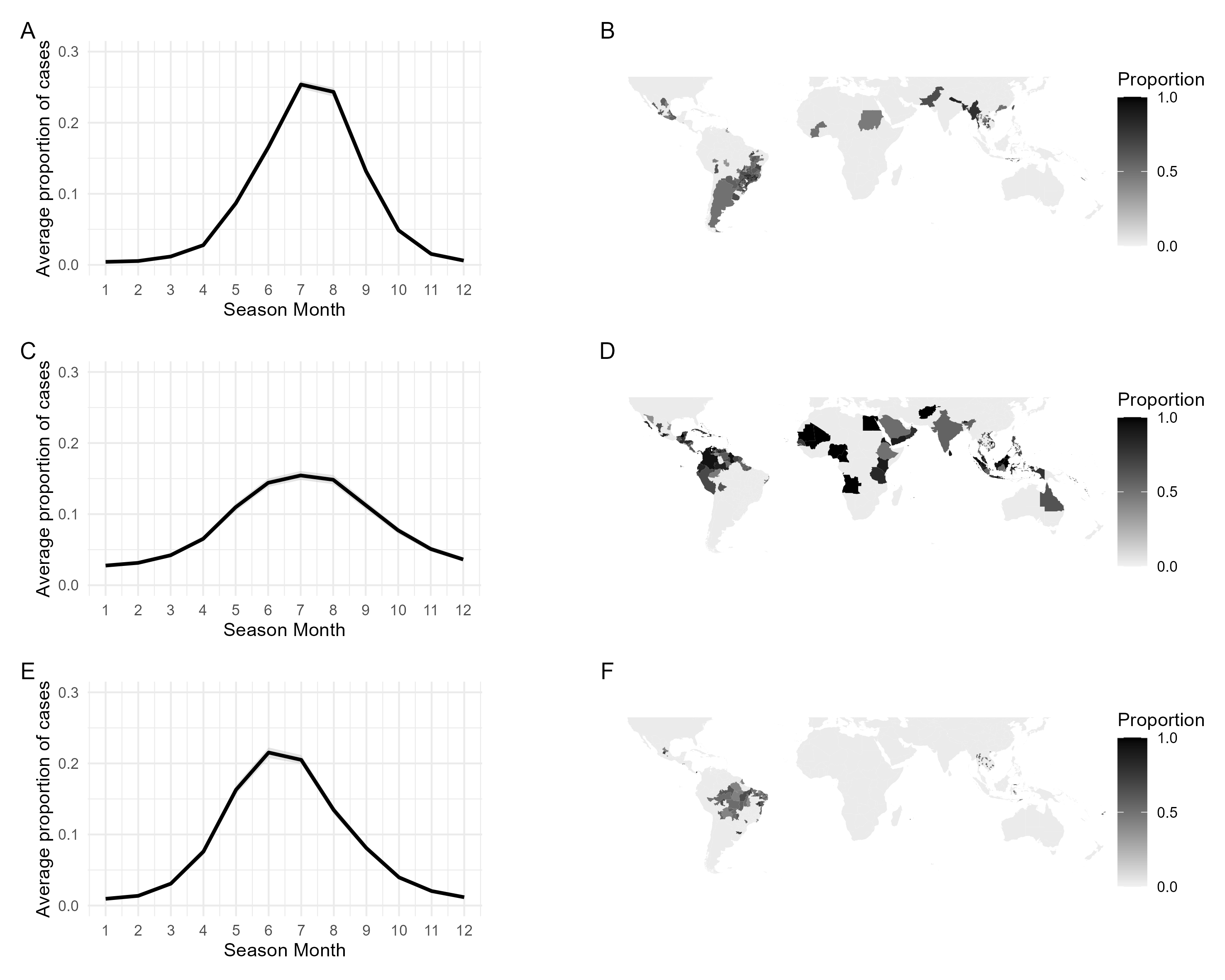

### Supplementary Figure 5

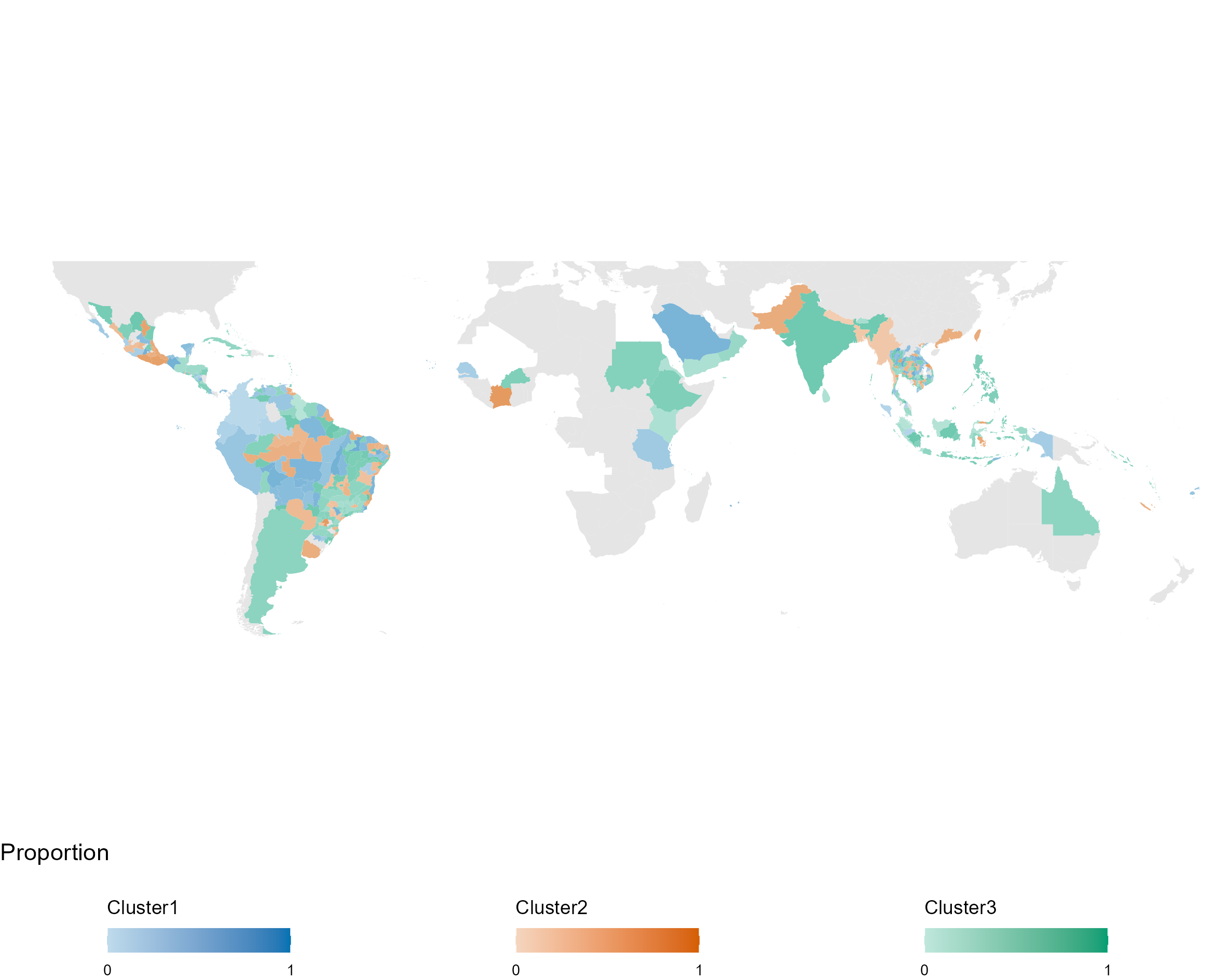

### Supplementary Figure 6

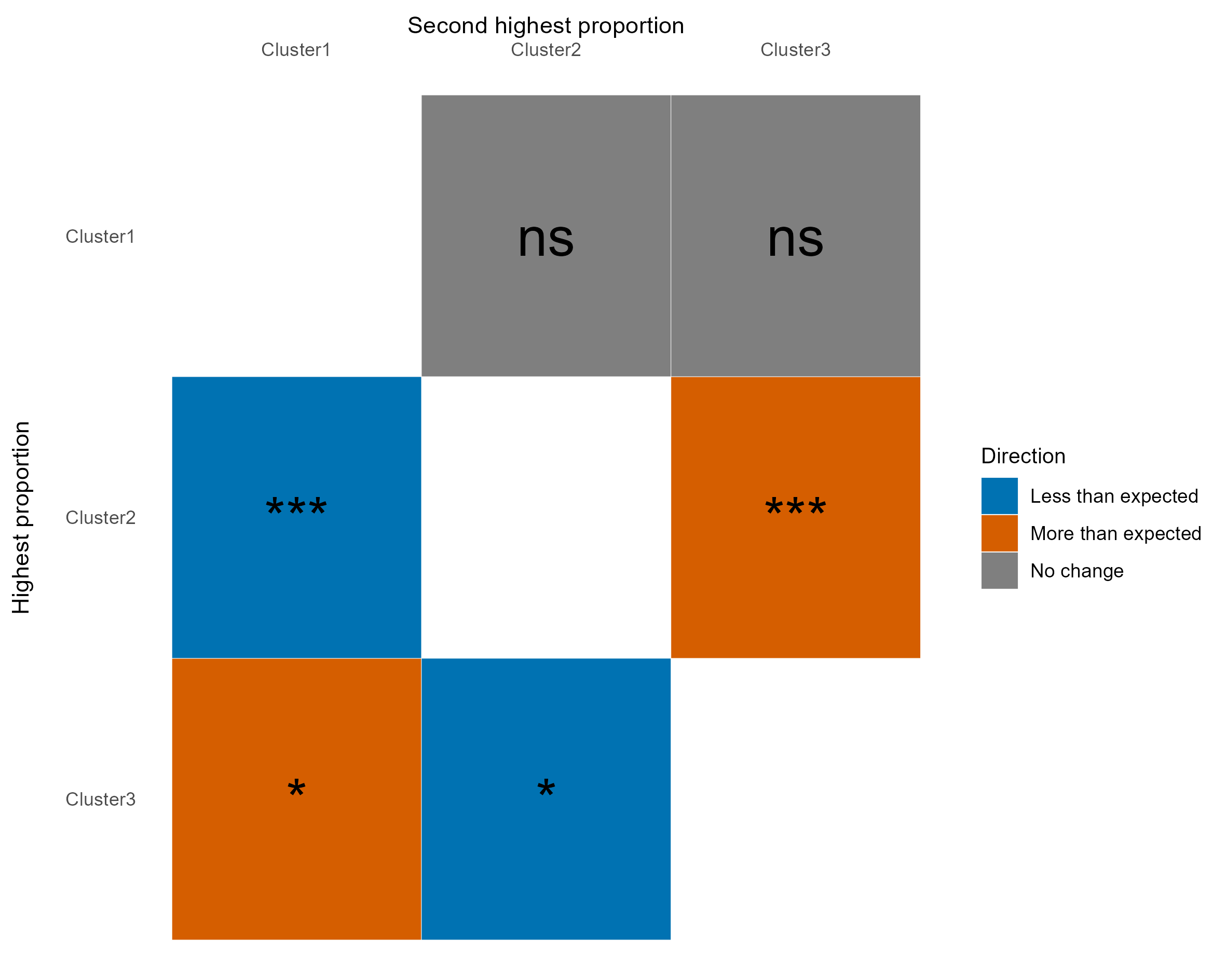

### Supplementary Figure 7

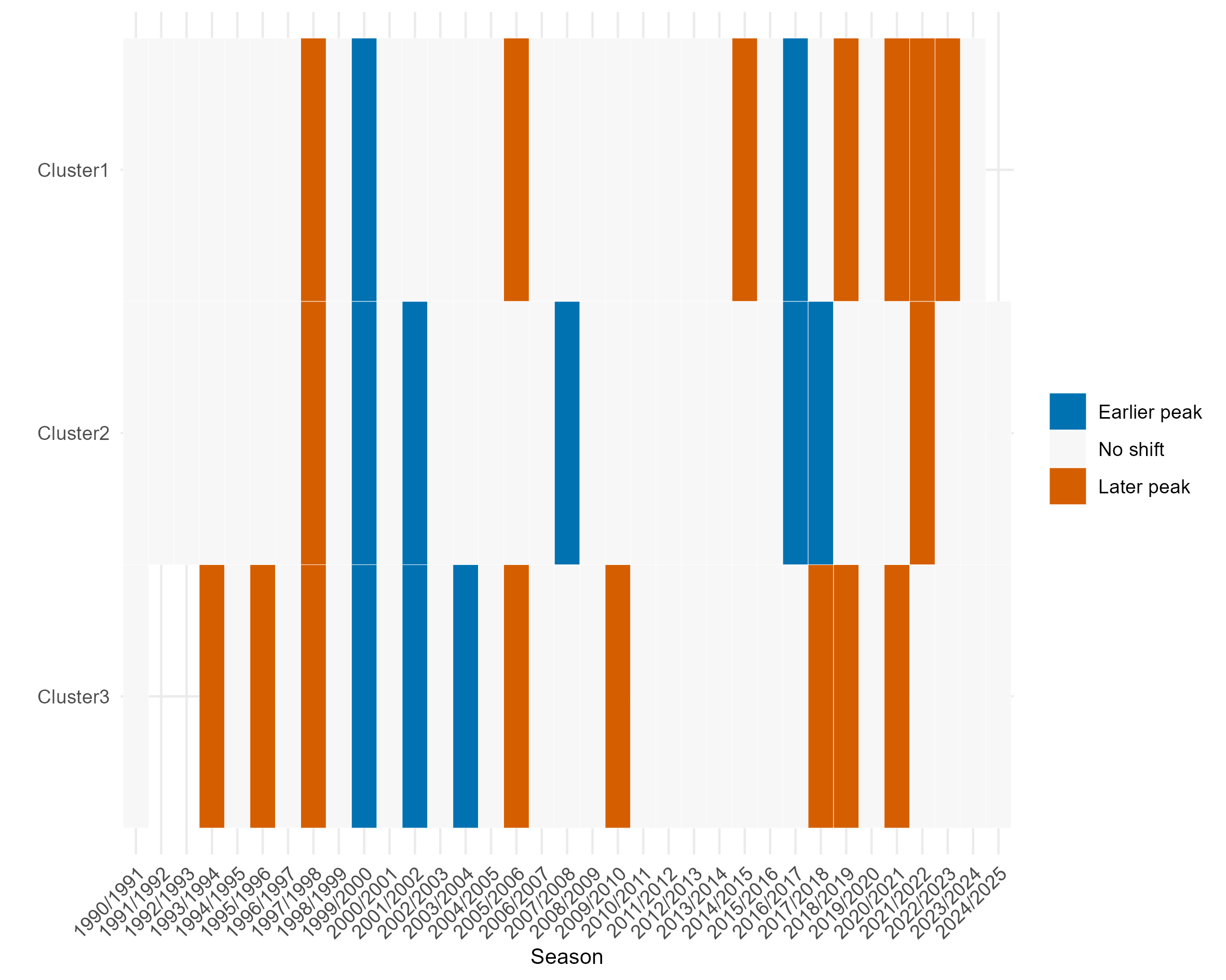

### Supplementary Table 6

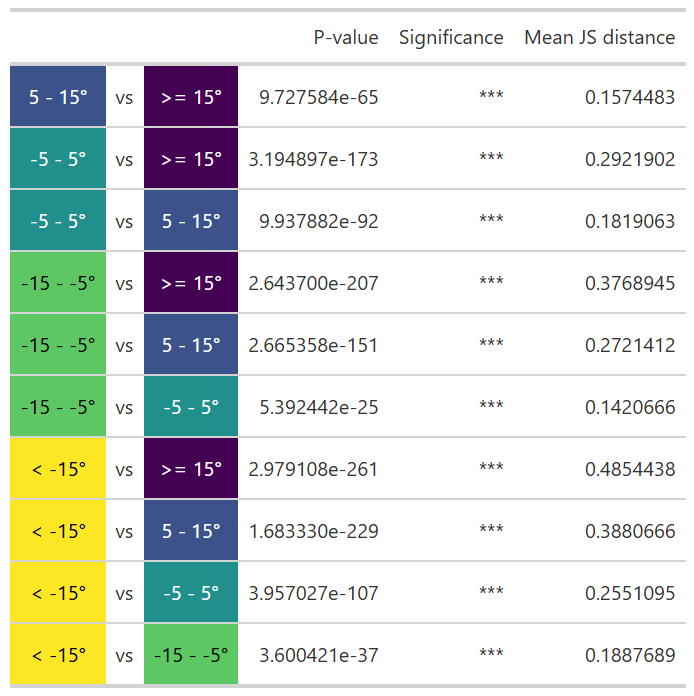
